# Chikungunya virus antepartum transmission and abnormal infant outcomes in Nigeria

**DOI:** 10.1101/2023.08.05.23293675

**Authors:** Atiene S. Sagay, Szu-Chia Hsieh, Yu-Ching Dai, Charlotte Ajeong Chang, Jerry Ogwuche, Olukemi O. Ige, Makshwar L. Kahansim, Beth Chaplin, Godwin Imade, Michael Elujoba, Michael Paul, Donald J. Hamel, Hideki Furuya, Ricardo Khoury, Viviane Sampaio Boaventura, Laíse de Moraes, Phyllis J. Kanki, Wei-Kung Wang

**Author notes:** Correspondence: Wei-Kung Wang and Phyllis J. Kanki. These authors contributed equally (Atiene S. Sagay, Szu-Chia Hsieh, Yu-Ching Dai).

## Abstract

Chikungunya virus (CHIKV) has become a global public health concern since the reemergence of the Indian Ocean lineage and expansion of the Asian genotype. CHIKV infection causes acute febrile illness, rash, and arthralgia and during pregnancy may affect both mothers and infants.

The mother-to-child transmission (MTCT) of CHIKV in Africa remains understudied. We screened 1006 pregnant women at two clinics in Nigeria between 2019 and 2022 and investigated the prevalence and MTCT of CHIKV. Of the 1006, 119 tested positive for CHIKV IgM, of which 36 underwent detailed laboratory tests. While none of the IgM reactive samples were RT-PCR positive, 14 symptomatic pregnant women were confirmed by CHIKV neutralization test. Twelve babies were followed with 8 normal and 4 abnormal outcomes, including stillbirth, cleft lip/palate with microcephaly, preterm delivery, polydactyly with sepsis and jaundice. CHIKV IgM testing identified 3 antepartum transmissions, further studies will determine its impact in antepartum infection.

## Introduction

Chikungunya virus (CHIKV), a member of the genus *Alphavirus* of the family *Togaviridae*, is transmitted by *Aedes aegypti* and *Aedes albopictus* mosquitoes. Prior to 2005, there were 3 distinct genotypes of CHIKV: the East-Central-South African (ECSA), West African and Asian genotypes in different geographic locations (*1–3*). The ECSA genotype spread from East Africa to the Indian Ocean and Asia between 2004-2007, known as the Indian Ocean lineage (IOL). In addition, the Asian genotype expanded in late 2013, the so-called the Asian-Western Hemisphere lineage, leading to its spread in the Americas with over 1.3 million suspected cases by April 2015 (*1–3*). It has been reported that autochthonous transmission of CHIKV occurs in 114 countries and territories over the tropical and sub-tropical areas of Africa, Asia, Oceania, the Americas, and Europe, where greater than three quarters of the world’s population reside (*1–3*).

CHIKV in Africa is maintained in an enzootic cycle involving non-human primates and sylvatic *Aedes* mosquitoes; sporadic cases or outbreaks tend to be small-scaled and associated with spillover from the natural reservoir vectors or environmental conditions such as increased rainfall (*4–6*). Recently, an increase in larger-scale outbreaks associated with urban areas of Africa have been reported (*2,7,8*).

CHIKV infection causes acute febrile illness, rash and arthralgia and may affect both pregnancy and infant outcomes (*9.10*). During the Reunion Island outbreak in 2005, the prospective study of CHIKV infection in pregnancy described mother-to-child transmission (MTCT) was observed in viremic mothers during the intrapartum period (within 2 days prior to and after delivery) with a transmission rate of 48.7%, leading to neonatal disease with sepsis-like illness, encephalitis, convulsions, and death (*11–13*). The study also described 3 exceptional cases of early fetal deaths (before 22 weeks) out of 678 antepartum CHIKV infections which were attributed to intrauterine CHIKV infection. As CHIKV spread from Asia to the Americas between 2007-2014, perinatal transmission of CHIKV was reported in India, Colombia, Brazil, and Curacao (*14–17*). Meta-analysis of cohort studies from Asia and Latin America revealed a MTCT rate of 15.5% with symptomatic neonatal disease attributed to intrapartum infection (*10,18,19*). However, the MTCT of African strains of CHIKV remains poorly understood.

In this study, we screened 1006 pregnant women at two antenatal clinics in Jos, north-central Nigeria from April 2019 to January 2022. We investigated the prevalence of CHIKV infection among pregnant women, transmission, and disease outcome in infants.

## Methods

### Nigeria cohort and human samples

Between April 1, 2019 and January 31, 2022, pregnant women aged ≥18 years, who attended the antenatal clinics at Jos University Teaching Hospital (JUTH) and Our Lady of Apostles Hospital (OLA) in Jos, north-central Nigeria, and presented with fever (≥37.5**°**C) or other symptoms (rash, headache, arthralgia, conjunctivitis, or myalgia) in the past 3 days were recruited with informed consent for screening by the Chembio DPP**^®^**ZCD IgM/IgG rapid test (Chembio Diagnostics Inc. Medford, NY). For every four symptomatic women, one asymptomatic woman with gestational age (GA) within two weeks of any of the four symptomatic women was also recruited for screening. All ZIKV, DENV or CHIKV rapid test IgM and IgM/IgG reactive women were recruited with informed consent for the prospective observational study as described (*20*). The pregnant women with CHIKV infection (CHIKV IgM+ and/or IgG+) and their infants were the focus of this study. The intrapartum period was defined as within 2 days prior to or after delivery (*11*); the remaining antepartum period included the first, second and third trimesters as defined by GA <12 wk, ≥12 wk to <28 wk, and ≥28 wk, respectively.

To evaluate the performance of the Chembio DPP**^®^** ZCD IgM/IgG rapid test, convalescent-phase sera from RT-PCR-confirmed CHIKV cases (n=22) in Brazil served as CHIKV-positive reference samples (*21*) and CHIKV-negative reference samples (n=27) were from a dengue seroprevalence study in Taiwan (*22*), a CHIKV non-endemic country; both were confirmed by CHIKV pseudovirus neutralization test (NT). Convalescent-phase serum/plasma samples from RT-PCR-confirmed DENV (n=35) and ZIKV (n=42) cases were also included in the analysis (Table S1) (*23*).

The study of coded serum or plasma samples was approved by the Institutional Review Boards (IRB) of the Harvard Longwood Campus (IRB# 17-0654), University of Jos (IRB# 127/XIX/5940), and University of Hawaii at Manoa (IRB# 2021-00044).

### Isolation of viral RNA and RT-PCR

Viral RNA was isolated from sera using the QIAamp viral RNA mini kit (Qiagen) and subjected to cDNA synthesis using the SuperScript™ III first-strand synthesis kit (Thermo Scientific); an aliquot of cDNA was subjected to nested PCR targeting a conserved region of the NSP2 gene of CHIKV (*24*). The RT-PCR products were electrophoresed through 2% agarose gel, and bands with the predicted size were purified for sequencing. The primers were CHIKV-F (5’-TCAATATGATGCAGATGAAAGT-3’, position 2541-2562), CHIKV-inner R (5’-GTCACAGGCA GTGTACACC -3’, position 2616-2634) and CHIKV-outer R (5’-GCAACGABGA CACAATGGC-3’, position 2536-2654). The size of the 1^st^ round and 2^nd^ round products were 113 bp and 93 bp, respectively. The PCR protocol will be provided upon request.

### IgM and IgG enzyme-linked immunosorbent assays (ELISAs) using DENV and ZIKV mutated virus like particles (VLP)

The plasmids expressing envelope protein fusion loop (FL)-mutated DENV1 VLP (Hawaii strain) and FL- and BC loop (BCL)-mutated ZIKV VLP (PRVABC59 strain), generation of VLP, and IgM and IgG ELISA using DENV FL-VLP or ZIKV FL/BCL VLP have been described previously (*25*). The sensitivity/specificity of DENV FL-VLP and ZIKV FL/BCL VLP IgG ELISAs was 100/93.3% and 100/83.3%, respectively; and that of DENV FL-VLP and ZIKV FL/BCL VLP IgM ELISAs was 75/95.8% and 90/99.3%, respectively.

### Microneutralization test

Microneutralization test was performed as described previously (*26,27*). Briefly, flat-bottom 96-well plates were seeded with Vero cells (3×10^4^ cells per well) 24 h prior to infection. Fourfold serial dilutions of serum (starting from 1:10 final dilution) were mixed with 50 focus-forming units of DENV1 (Hawaii strain), DENV2 (NGC strain), DENV3 (CH53489), DENV4 (H241 strain), or ZIKV (PRVABC59 strain) at 37°C for 1 h. The mixtures were added to each well followed by incubation for 48 h (except 70 h for DENV1), removal of medium, and fixation as described previously (*26,27*). After adding a murine monoclonal antibody 4G2 and secondary antibody mixture (IRDye 800CW-conjugated goat anti-mouse IgG at 1:10,000 and DRAQ5 fluorescent probe at 1:10,000), the signal (800 nm/700 nm fluorescence) was detected using a Li-Cor Odyssey Classic imaging system (Li-Cor Biosciences, Lincoln, NE) and analyzed using Image Studio software to determine percent neutralization at different concentrations and NT_90_ as described previously (*26,27*).

### Cell lines and plasmids

Human hepatoma Huh7 cells were obtained from Health Science Research Resources Bank, Japan Health Sciences Foundation (Osaka, Japan). HEK-293T cells were obtained from the American Type Culture Collection (Manassas, VA). Plasmids pNL4-3 R-E-miRFP, which contains the monomeric infrared fluorescent protein (miRFP) gene replacing the Luc gene of an *env*-defective HIV-1 reporter construct pNL4-3.Luc.R-E-, has been described previously (*28*). The E3-E2-6K-E1 genes of CHIKV (2955 bp) and Mayaro virus (MAYV) (2952 bp) were PCR amplified from cDNA derived from RNA of CHIKV (H20235 strain) and MAYV (TRVL strain), respectively, (BEI Resources, Manassas, VA), and cloned into pCB vector (by NotI and XhoI sites) to generate plasmids CHIKV and MAYV. All plasmids were confirmed by sequencing of the entire E3-E2-6K-E1 gene insert and verified for expression by transfection and Western blot analysis (*28*).

### Generation of CHIKV and MAYV pseudoviruses

To generate pseudoviruses, HEK-293T cells were seeded in 10-cm dish one day before transfection, co-transfected with pNL4-3 R-E-miRFP (12 µg) and plasmids CHIKV (12 µg) or MAYV (4 µg) using lipofectamine 2000 and incubated with DMEM media containing 10% FBS (*28*). The supernatants were collected at 72 h post transfection, followed by low-speed centrifugation at 300x g for 10 min, aliquoted and stored at -80°C. To titrate each pseudovirus, 3-fold serially diluted supernatants were inoculated to Huh7 cells by spin infection (*28*); miRFP signals were quantitated at 72 h post-infection, and the amount of pseudovirus that resulted in miRFP signals 10 times higher than the mock-infected wells was used for the NT.

### CHIKV and MAYV pseudovirus NT

Huh7 cells (2 x 10^4^ cells/well) were seeded onto 96-well plates one day before infection. Pseudovirus (CHIKV or MAYV) was mixed with 4-fold serial dilutions of plasma at 1:1 ratio, incubated at 37°C for 1 h, and added to each well for spin infection. At 72 h, the plates were scanned by Li-Cor Odyssey imager (*28*). The % of infection at different plasma dilutions (from 1:10 to 1:10,240 dilutions) were calculated by the formula (intensity of serum+pseudovirus – intensity of media only)/(intensity of pseudovirus only – intensity of media only) x 100. The % neutralization=100 – % of infection (*28*). NT_50_ titer was the plasma dilution that reached 50% neutralization using 4-parameter nonlinear regression analysis (GraphPad 6.0, Boston, MA) (Figure 1). NT_50_ titer<10 was arbitrarily assigned as 5.

**Figure 1.**
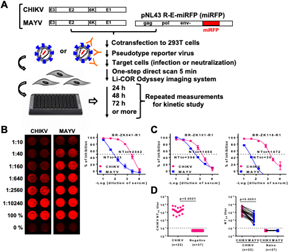
Generation of CHIKV and MAYV pseudoviruses and NT. (A) Schematic drawing of plasmids expressing E2 and E1 proteins (E3-E2-6K-E1 genes) of CHIKV or MAYV and co-transfection with pNL43 R-E-miRFP (miRFP) to generate CHIKV or MAYV pseudoviruses containing miRFP reporter, infection of target cells and one-step imagining. (B) NTs based on CHIKV and MAYV pseudoviruses with miRFP reporter. The miRFP signals (left) and neutralization curves and NT_50_ titers (right) to CHIKV and MAYV pseudoviruses at 72 h post-infection in Huh7 cells of convalescent-phase serum sample from an RT-PCT-confirmed CHIKV case from Brazil. (C) Neutralization curves and NT_50_ titers to CHIKV and MAYV pseudoviruses of other 2 confirmed CHIKV cases. (D) NT_50_ titers to CHIKV and MAYV pseudoviruses of 22 confirmed CHIKV cases and 27 CHIKV-negative individuals. The two-tailed Mann-Whitney (left) and Wilcoxon signed-rank (right) tests (GraphPad 6.0).

### Statistical analysis

The two-tailed Fisher’s exact test (Pearson Chi-square test) and two-tailed Mann-Whitney test were used to compare qualitative and quantitative variables, respectively, between two groups; the two-tailed Wilcoxon signed-rank test paired data (GraphPad 6.0, Boston, MA).

## Results

### Detection of CHIKV infection

The Chembio DPP**^®^** ZCD IgM/IgG rapid test was used to detect CHIKV IgM and IgG in consenting pregnant women at the antenatal clinics of JUTH and OLA. To assess the sensitivity/specificity of the CHIKV IgM and IgG assays, we first tested with a panel of convalescent-phase samples from RT-PCR-confirmed CHIKV, DENV and ZIKV cases and CHIKV-negative samples from a dengue seroprevalence study in Taiwan (*21–23*). The sensitivity/specificity of the CHIKV IgM and IgG assays was 90.9/95.2% and 100/100%, respectively (Table S1).

Between April 2019 and January 2022, 787 febrile or otherwise symptomatic and 219 asymptomatic women were screened. A third of women (312/1006, 31.0%) were reactive to ZIKV, DENV, CHIKV or some combination. CHIKV IgG was detected in 120/1006, suggesting a seroprevalence of 11.9%, and CHIKV IgM in 119/1006 (11.8%), suggesting acute or recent CHIKV infection. Among symptomatic women, headache was the most common symptom followed by fever and fatigue. Unlike similar studies in the Americas, rash and arthralgia were not commonly reported. The prevalence of CHIKV IgM was higher in asymptomatic women (41/219, 18.7%) compared to symptomatic women (78/787, 9.9%). CHIKV RT-PCR was performed but none of the CHIKV IgM reactive samples tested positive.

### Confirmation of CHIKV infection by NT

We further generated CHIKV pseudovirus based on a lentivirus vector with an miRFP reporter and developed a CHIKV NT (Figure 1) (*28*). When testing with a panel of convalescent-phase sera from RT-PCR-confirmed CHIKV cases in Brazil, we found high NT_50_ titers (289–6867) to CHIKV (Figure 1). As a comparison, a MAYV pseudovirus NT was also performed and lower NT_50_ titers (30–1522) to MAYV were observed; the ratio of CHIKV NT_50_ titers to MAYV NT_50_ titers were higher than 2.5, which was consistent with two recent reports (*29,30*).

We next used the CHIKV NT to examine 36 samples, collected during the first two years of the study, from 119 pregnant women with acute or recent CHIKV infection (either IgM+IgG+ or IgM+IgG−). Of the 36 samples, we found 18 with detectable neutralizing antibodies to CHIKV (16 from the IgM+IgG+ subgroup and 2 from the IgM+IgG− subgroup) (Table 1, Figure 2A). This is in agreement with the observation of distinct antibody patterns and the presence of an IgM+IgG− period in some individuals following CHIKV infection (*31*).

**Figure 2.**
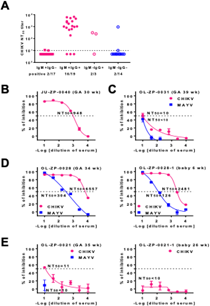
CHIKV pseudoviruses NT in Nigeria pregnant women. (A) CHIKV NT_50_ titers in different subgroups based on CHIKV IgM/IgG results using Chembio DPP^®^ ZCD IgM/IgG rapid test. (B-E) Neutralization curves and NT_50_ titers to CHIKV and MAYV pseudoviruses of 4 pregnant women with acute CHIKV infection and abnormal infant outcomes. Data are means and standard deviations of duplicates from one experiment.

**Table 1.** Serological evidence of CHIKV infection during pregnancy and infant outcomes

| CHIKV Serological tests* | Pregnant women<br>n=36 | Symptomatic pregnant women<br>n=30 | Infants follow-up<br>n=26 | Infant outcomes |  |
| --- | --- | --- | --- | --- | --- |
|  |  |  |  | normal<br>n=19 | abnormal<br>n=7 |
| IgM+IgG- | 17 | 15 |  |  |  |
| NT+ | 2 | 2 | 2 | 0 | 1 multiple congenital anomalies (cleft lip/palate/microcephaly)<br>1 polydactyly/sepsis/jaundice |
| NT- | 15 | 13 | 11 | 8 | 1 stillbirth<br>1 stillbirth and preterm<br>1 multiple congenital anomalies (hypotonia/seizure/microcephaly) |
| IgM+IgG+ | 19 | 15 |  |  |  |
| NT+ | 16 | 12 | 10 | 8 (2 antepartum transmission) | 1 stillbirth<br>1 preterm (antepartum transmission) |
| NT- | 3 | 3 | 3 | 3 | 0 |

\* Chembio DPP® ZCD IgM/IgG rapid test was performed to detect CHIKV IgM and IgG, followed by CHIKV pseudovirus neutralization test (NT).

### Infant outcomes of antepartum CHIKV infection

Of the 36 pregnant women with CHIKV IgM reactivity (either IgM+IgG+ or IgM+IgG−), 30 were symptomatic suggesting acute CHIKV infection. Of the 30 women, 26 babies were followed up and 19 had normal and 7 had abnormal outcomes including three stillbirths, two multiple congenital anomalies, one polydactyly with sepsis and jaundice, and one preterm (Table 1). In the subset of 14 acute CHIKV infections confirmed by NT, 12 babies were followed with 8 normal and 4 abnormal outcomes.

The time course of 4 mothers with acute CHIKV infection confirmed by NT (Figures 2B-2E) and their babies with abnormal outcomes was summarized in Figures 3A-3D. Notably, babies with multiple congenital anomalies may have lower odds of survival. Case OL-ZP-0031 was enrolled at 39 wk with CHIKV IgM+/IgG- and NT_50_ titer of 10 to CHIKV; she gave birth to a baby with multiple congenital anomalies including cleft lip, palate and microcephaly who passed away in 2 days (Figure 3A). Additionally, past flavivirus infection was confirmed by NT. Case OL-ZP-0028 was enrolled at 33 wk with CHIKV IgM+/IgG+ and high NT_50_ titer (of 6657) to CHIKV; due to ZIKV ELISA IgG+, NT was performed and showed NT_90_ titer of 51 to ZIKV and <10 to DENV1-4, suggesting past ZIKV infection (Figure 3D).

**Figure 3.**
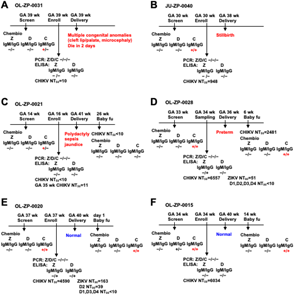
Time course of pregnant women who had acute CHIKV infection confirmed by NT and gave birth to babies with abnormal outcomes or antepartum transmission. (A-D) Time course of 4 pregnant women with acute CHIKV infection and abnormal infant outcomes. Multiple congenital anomalies with cleft lip/palate and microcephaly (A), stillbirths (B), polydactyly/sepsis/ jaundice (C), and preterm (D). (D, E-F) Time course of 3 pregnant women with acute CHIKV infection and antepartum transmission of CHIKV to fetus. GA, gestational age; wk, weeks; Z, Zika virus; D, dengue virus; C, chikungunya virus. CHIKV IgM-reactive infants highlighted in red.

### Antepartum transmission of CHIKV

Of the 26 CHIKV infections among symptomatic pregnant women with babies followed, one was detected in the first trimester, 8 second trimester and 17 third trimester (Table 2). We further examined the babies’ samples to assess the possibility of antepartum transmission; of the 19 available samples from babies followed, 3 were CHIKV IgM reactive (all in the third trimester with two normal and one preterm outcomes), suggesting antepartum transmission of CHIKV (Table 1). The time course of 3 pregnant women with antepartum CHIKV infection and transmission to their babies were summarized in Figures 3D and 3E-3F. Case OL-ZP-0028 gave birth to a preterm baby at 36 wk; the baby at 6 wk (the first follow-up) showed CHIKV IgM+/IgG+ (Figure 3D). Another past flavivirus infection confirmed by NT was noted in case OL-ZP-0020, who was enrolled at 37 wk with CHIKV IgM+/IgG+ and high NT_50_ titer to CHIKV; due to ZIKV and DENV ELISAs IgG+, NT to ZIKV and DENV was performed and showed NT_90_ titers of 163 to ZIKV, 39 to DENV2 and <10 to DENV1, 3 and 4, suggesting past ZIKV and DENV infection. She gave birth to a normal baby at 40 wk; the baby followed up at day 1 showed CHIKV IgM+/IgG+ (Figure 3E).

**Table 2.** Time of CHIKV infection during pregnancy and infant outcomes

| CHIKV Serological tests* | Pregnant women<br>n=36 | Symptomatic pregnant women<br>n=30 | Infants follow-up<br>n=26 | CHIKV infection during pregnancy |  |  |  |
| --- | --- | --- | --- | --- | --- | --- | --- |
|  |  |  |  | 1<sup>st</sup> trimester<br>n=1 | 2<sup>nd</sup> trimester<br>n=8 | 3<sup>rd</sup> trimester prior to labor<br>n=17 | intrapartum<br>n=0 |
| IgM+ IgG- | 17 | 15 |  |  |  |  |  |
| NT+ | 2 | 2 | 2 | 0 | 1 (†) | 1 (¶) | 0 |
| NT- | 15 | 13 | 11 | 1 | 4 | 6 (\$‡§) | 0 |
| IgM+ IgG+ | 19 | 15 |  |  |  |  |  |
| NT+ | 16 | 12 | 10 | 0 | 1 | 9 (‡#) | 0 |
| NT- | 3 | 3 | 3 | 0 | 2 | 1 | 0 |

\* Chembio DPP® ZCD IgM/IgG rapid test was performed to detect CHIKV IgM and IgG, followed by CHIKV pseudovirus neutralization test (NT).
Infants with abnormal outcomes are shown in parenthesis:
† polydactyly/sepsis/jaundice.
¶ multiple congenital anomalies (cleft lip/palate/microcephaly).
\$ multiple congenital anomalies (hypotonia/seizure/microcephaly).
‡ stillbirth.
§ stillbirth and preterm. The time course of 3 pregnant women with acute CHIKV infection not confirmed by NT and their infants with abnormal outcomes was summarized in Appendix Figure 1.

## Discussion

In this study, we examined CHIKV infection during pregnancy in a non-outbreak setting in Nigeria and reported antepartum transmission of CHIKV and association of antepartum CHIKV infection with abnormal infant outcomes.

Based on previous reports from the 2005 Reunion Island outbreak that MTCT of CHIKV occurred predominantly during the intrapartum period, subsequent studies of CHIKV infection during pregnancy primarily focused on the perinatal period and confirmed the contribution of intrapartum CHIKV infection to the MTCT of CHIKV with IOL and Asian genotypes in Asia and Latin America (*14–19*). Our finding of CHIKV transmission to 3 infants out of 19 (15.8%) pregnant women with antepartum CHIKV infection was unexpected compared with the Reunion Island study, in which 3 abortions out of 678 (0.4%) antepartum CHIKV infections were attributed to in utero CHIKV infection. The higher rates of antepartum CHIKV MTCT might be attributed to pathogenesis of endemic infection and/or CHIKV strain differences. Furthermore, we reported 7 abnormal infant outcomes including three stillbirths, two multiple congenital anomalies, one polydactyly with sepsis and jaundice, and one preterm associated with 26 antepartum CHIKV infections (26.9%). The 7 abnormal infant outcomes were a subset of 17 abnormal infant outcomes associated with arbovirus (ZIKV,DENV, and CHIKV) infection during pregnancy, in which birth registry data were compared (*20*). Notably, another study of the CHIKV outbreak in Colombo, Sri Lanka in 2006-2007 reported transmission of CHIKV to 6 infants (based on newborn IgM positivity) out of 46 (13.0 %) pregnant women with antepartum CHIKV infection and 10 (21.7%) abnormal outcomes including two abortions, one intrauterine death, one preterm, five congenital heart diseases (atrial septal defect, patent ductus arteriosus and persistent foramen ovalis) and one intrauterine growth retardation (*32*). As the Sri Lanka outbreak was likely to be caused by the IOL, the difference between our findings and those from the Reunion Island outbreak could not be explained by different CHIKV genotypes alone.

A recent review of 13 CHIKV outbreaks in Africa from 1999 to 2020 revealed that viral lineage was identified in 8 outbreaks; all were associated with ECSA genotype (which the IOL originated from) except one outbreak of West African genotype in Kedougou, Senegal during 2009-2010, in which 14 out of 144 acute sera at 5 health care facilities were confirmed by CHIKV RT-PCR and 6 CHIKV IgM+ cases were identified from 1409 sera collected (*2,5,6*). The timing of sample collection after symptom onset differed between studies and may account for the difference in detection by RT-PCR.

After CHIKV infection, individuals develop an IgM response starting from 3 to 8 days post-symptom onset, followed by an IgG response at 7 to 14 days (*33–35*). A recent study reported 5 distinct antibody patterns (IgM−IgG−/NT−, IgM+IgG−/NT−, IgM+IgG−/NT+, IgM+IgG+/NT− and IgM+IgG+/NT+) during acute febrile phase and the presence of NT or IgG antibody was associated with protection against developing chronic arthritis in the future, which was supported by two studies of outbreaks in India (2010-3 and 2014-6) (*31,36*). Although most individuals of the IgM−IgG− and IgM+ IgG− subgroups developed IgG and NT antibodies at the convalescent-phase, this may explain some IgM+IgG− and NT− samples observed in our study.

The proportion of inapparent CHIKV infection has been reported to be 3%–25%, corresponding to a symptomatic to inapparent ratio of 1:0.03 to 1:0.33 (*2,37*). Recent studies of the outbreak in Nicaragua reported a higher proportion of inapparent infection with a symptomatic to inapparent ratio of 1:0.65 to 1:1.20, which varies between lineages and epidemics (*38,39*). Our findings of a symptomatic to inapparent ratio of 1:2 is unexpected, which may be related to viral lineage differences and the non-outbreak setting.

Conventional plaque-reduction neutralization test (PRNT) for CHIKV requires a biosafety level 3 laboratory and is labor intensive and time-consuming (*35*). Several NTs based on different CHIKV pseudoviruses, replicon particles or genetically modified infectious clones containing luciferase or green fluorescent protein as reporters have been developed (*40–44*). Compared with live virus PRNT, our CHIKV pseudovirus NT can be performed in a biosafety level 2 laboratory and requires fewer plates (one 96-well plate for 6 samples in duplicates vs. eighteen 6-well plates for PRNT), less time (72 h vs. >4 days for PRNT), and less sample volume (20 µL vs. 100 µL for PRNT in duplicates, starting from serum at 1:10 final dilution). Compared with CHIKV pseudovirues using luciferase reporter, our CHIKV pseudovirus employed miRFP reporter which can be quantified by one step of direct imaging without multiple laborious steps (5 min scan vs. ∼60 min for a 96-well plate) (Figure 1). Moreover, the same plate can be quantified multiple times for kinetic study without generating numerous replicates. Compared with GFP reporter, miRFP has minimal autofluorescence and can be quantified by either direct imaging or flow cytometry to determine the percentage of positive cells. Together, these features suggest our CHIKV pseudovirus with miRFP reporter is a simple, practical, and cost-effective tool for neutralization.

There are several limitations to our study. First, the sample size of pregnant women with CHIKV infection (IgM+) analyzed (n=36) was small; future studies involving larger sample size are needed to validate these observations. There was no difference in age, ethnic groups, trimesters, and ratio of symptomatic vs. inapparent infection between the 36 pregnant women analyzed and the 83 pregnant women not analyzed in this study (Pearson Chi-square test), except that the 36 women were enrolled exclusively during the first two years of the study. Therefore, the analysis of this subset was unlikely to affect our results. Second, although the babies’ samples after delivery or the first follow-up were assessed to identify antepartum transmission, the possibility of neonate CHIKV infection cannot be completely ruled out. For example, the first available follow-up sample for the baby of case OL-ZP-0015 was at 14 wk (Figure 3F); excluding this case resulted in an antepartum transmission rate of 10.5% (2/19), which was still higher than that (0.4%) reported in the Reunion Island study. Third, NT to rule out O’nyong-nyong virus (ONNV), a closely related alphavirus which caused intermittent yet explosive outbreaks in East Africa, was not performed (*35,45*). Despite previous report of the unidirectional antigenic relationships between the two viruses, some sera still cannot be distinguished by NT (*46–49*). Since the ONNV outbreak involving both the East and West Africa in 1959-1962, ONNV has been apparently silent for 35 years until 1996-1997 when another outbreak in East Africa (Uganda and Kenya and Tanzania) took place (*45,48–50*). In Nigeria, only 3 confirmed-ONNV cases (by virus isolation) were reported in 1966 and 1969, suggesting the possibility of ONNV infection among the participants in Nigeria is remote (*45*).

CHIKV was first described in Africa, yet subsequent to the Reunion Island outbreak the epidemiology and pathogenesis in the continent has been infrequently studied. Our study provides new and convenient diagnostic tools for future study. In addition, we describe significant asymptomatic infection and abnormal pregnancy and infant outcomes associated with antepartum CHIKV infection. Future studies are needed to further characterize the risk of antepartum CHIKV infection to pregnant women and their infants in Africa.

## Supplementary Data

Appendix Table 1 and Figure 1.

## Data Availability

All data produced in the present study are available upon reasonable request to the authors

## Acknowledgments

We thank the pregnant women at the JUTH and OLA clinics that participated in this study to advance our knowledge of arbovirus infection. We also thank the clinical, laboratory and data management teams at JUTH and OLA whose efforts enabled this work. This work was supported by grants R21AI137840 (Kanki) and R01AI149502-03 (Wang) from the National Institute of Allergy and Infectious Diseases, 5D43TWO10130 (Sagay) from the Fogarty International Center, NIH, and MedRes-2022-0000 0789 (Wang) from the Hawaii Community Foundation. The funders had no role in study design, data collection and analysis, decision to publish, or preparation of the manuscript.

## About the Author

Dr. Atiene Sagay is a Professor of Obstetrics and Gynecology at the University of Jos, Nigeria, his research centers on maternal health. Dr. Szu-Chia Hsieh was a Research Scientist at the Department of Tropical Medicine, Medical Microbiology and Pharmacology at the University of Hawaii at Manoa during this study and is currently relocated to the Graduate Institute of Anatomy and Cell Biology, College of Medicine, National Taiwan University; her research focuses on arboviruses. Ms. Yu-Ching Dai was a Research Assistant and currently Graduate student at the Department of Tropical Medicine, Medical Microbiology and Pharmacology at the University of Hawaii at Manoa; her research focuses on arboviruses.

## Notes

### Competing Interest Statement

The authors have declared no competing interest.

## References

1. Weaver SC, Lecuit M. Chikungunya virus and the global spread of a mosquito-borne disease. N Engl J Med. 2015;372:1231–9.

2. Bettis AA, L’Azou Jackson M, Yoon IK, Breugelmans JG, Goios A, Gubler DJ, et al. The global epidemiology of chikungunya from 1999 to 2020: A systematic literature review to inform the development and introduction of vaccines. PLoS Negl Trop Dis. 2022;16:e0010069.

3. Puntasecca CJ, King CH, LaBeaud AD. Measuring the global burden of chikungunya and Zika viruses: A systematic review. PLoS Negl Trop Dis. 2021;15:e0009055.

4. Pezzi L, Reusken CB, Weaver SC, Drexler JF, Busch M, LaBeaud AD, et al. GloPID-R report on Chikungunya, O’nyong-nyong and Mayaro virus, part I: Biological diagnostics. Antiviral Res. 2019;166:66–81.

5. Sow A, Faye O, Diallo M, Diallo D, Chen R, Faye O, et al. Chikungunya Outbreak in Kedougou, Southeastern Senegal in 2009-2010. Open Forum Infect Dis. 2017;5:ofx259.

6. Althouse BM, Guerbois M, Cummings DAT, Diop OM, Faye O, Faye A, et al. Role of monkeys in the sylvatic cycle of chikungunya virus in Senegal. Nat Commun. 2018;9:1046.

7. Renault P, Solet JL, Sissoko D, Balleydier E, Larrieu S, Filleul L, et al. A major epidemic of chikungunya virus infection on Reunion Island, France, 2005-2006. Am J Trop Med Hyg. 2007;77:727–31.

8. Peyrefitte CN, Bessaud M, Pastorino BA, Gravier P, Plumet S, Merle OL, et al. Circulation of Chikungunya virus in Gabon, 2006-2007. J Med Virol. 2008;80:430–3.

9. Charlier C, Beaudoin MC, Couderc T, Lortholary O, Lecuit M. Arboviruses and pregnancy: maternal, fetal, and neonatal effects. Lancet Child Adolesc Health. 2017;1:134–46.

10. Contopoulos-Ioannidis D, Newman-Lindsay S, Chow C, LaBeaud AD. Mother-to-child transmission of chikungunya virus: A systematic review and meta-analysis. PLoS Negl Trop Dis. 2018;12:e0006510.

11. Gérardin P, Barau G, Michault A, Bintner M, Randrianaivo H, Choker G, et al. Multidisciplinary prospective study of mother-to-child chikungunya virus infections on the island of La Réunion. PLoS Med. 2008;5:e60.

12. Fritel X, Rollot O, Gerardin P, Gauzere BA, Bideault J, Lagarde L, et al. Chikungunya virus infection during pregnancy, Reunion, France, 2006. Emerg Infect Dis. 2010;16:418–25.

13. Ramful D, Sampériz S, Fritel X, Michault A, Jaffar-Bandjee MC, Rollot O, et al. Antibody kinetics in infants exposed to chikungunya virus infection during pregnancy reveals absence of congenital infection. J Infect Dis. 2014;209:1726–30.

14. Kumar S, Agrawal G, Wazir S, Kumar A, Dubey S, Balde M, et al. Experience of perinatal and neonatal chikungunya virus (Chikv) infection in a tertiary care neonatal centre during outbreak in North India in 2016: a case series. J Trop Pediatr. 2019;65:169–75.

15. Villamil-Gomez W, Alba-Silvera L, Menco-Ramos A, Gonzalez-Vergara A, Molinares-Palacios T, Barrios-Corrales M, et al. Congenital chikungunya virus infection in Sincelejo, Colombia: a case series. J Trop Pediatr. 2015;61:386-92.

16. Bandeira AC, Campos GS, Sardi SI, Rocha VF, Rocha GC. Neonatal encephalitis due to chikungunya vertical transmission: first report in Brazil. ID Cases. 2016;5:57–9.

17. van Enter BJD, Huibers MHW, van Rooij L, Steingrover R, van Hensbroek MB, Voigt RR, et al. Perinatal outcomes in vertically infected neonates during a chikungunya outbreak on the Island of Curacao. Am J Trop Med Hyg. 2018;99:1415–8.

18. Torres JR, Falleiros-Arlant LH, Dueñas L, Pleitez-Navarrete J, Salgado DM, Castillo JB. Congenital and perinatal complications of chikungunya fever: a Latin American experience. Int J Infect Dis. 2016;51:85–88.

19. Escobar M, Nieto AJ, Loaiza-Osorio S, Barona JS, Rosso F. Pregnant women hospitalized with chikungunya virus infection, Colombia, 2015. Emerg Infect Dis. 2017;23:1777–83.

20. Ogwuche J, Chang CA, Ige O, Sagay AS, Chaplin B, Kahansim ML, et al. Arbovirus surveillance in pregnant women in north-central Nigeria, 2019-2022. submitted. 2023.

21. de Moraes L, Cerqueira-Silva T, Nobrega V, Akrami K, Santos LA, Orge C, et al. A clinical scoring system to predict long-term arthralgia in chikungunya disease: A cohort study. PLoS Negl Trop Dis. 2020;14(7):e0008467.

22. Tsai JJ, Liu CK, Tsai WY, Liu LT, Tyson J, Tsai CY, et al. Seroprevalence of dengue in two districts of Kaohsiung city after the largest dengue outbreak in Taiwan since world war II. PLoS Negl Trop Dis 2018;12:e0006879.

23. Tyson J, Tsai WY, Tsai JJ, Brites C, Mässgård L, Youn HH, et al. Combination of non-structural protein 1-based enzyme-linked immunosorbent assays can detect and distinguish various dengue virus and Zika virus infections. J Clin Microbiol. 2019;57: e01464–18.

24. Waggoner JJ, Ballesteros G, Gresh L, Mohamed-Hadley A, Tellez Y, Sahoo MK, et al. Clinical evaluation of a single-reaction real-time RT-PCR for pan-dengue and chikungunya virus detection. J Clin Virol. 2016;78:57–61.

25. Tsai WY, Driesse K, Tsai JJ, Granat RS, Jenkins O, Hsieh SC, et al. Enzyme-linked immunosorbent assays using virus-like particles containing mutations of conserved residues on envelop protein can distinguish three flavivirus infections. Emerg Microbe Infect 2020;9:1722–32.

26. Tsai WY, Chen HL, Tsai JJ, Dejnirattisai W, Jumnainsong A, Mongkolsapaya J, et al. Potent neutralizing human monoclonal antibodies preferentially target mature dengue virus particles: implication for novel strategy of dengue vaccine. J Virol. 2018;92:e00556–18.

27. Herrera BB, Tsai WY, Brites C, Luz E, Pedroso C, Drexler JF, et al. T cell responses to nonstructural protein 3 distinguish infections by dengue and Zika viruses. mBio 2018;9: e00755–18.

28. Tsai WY, Ching LC, Hsieh SC, Melish ME, Nerriah VR, Wang WK. A real-time and high-throughput neutralization test based on SARS-CoV-2 pseudovirus containing monomeric infrared fluorescent protein as reporter. Emerg Microbe Infect. 2021;10:894–905.

29. Martins KA, Gregory MK, Valdez SM, Sprague TR, Encinales L, Pacheco N, et al. Neutralizing antibodies from convalescent chikungunya virus patients can cross-neutralize Mayaro and Una viruses. Am J Trop Med Hyg. 2019;100:1541–44.

30. Bopp NE, Jencks KJ, Siles C, Guevara C, Vilcarromero S, Fernández D, et al. Serological responses in patients infected with Mayaro virus and evaluation of cross-protective responses against chikungunya virus. Am J Trop Med Hyg. 2021;106:607–9.

31. Nayak K, Jain V, Kaur M, Khan N, Gottimukkala K, Aggarwal C, et al. Antibody response patterns in chikungunya febrile phase predict protection versus progression to chronic arthritis. JCI Insight. 2020;5:e130509.

32. Senanayake MP, Senanayake SM, Vidanage KK, Gunasena S, Lamabadusuriya SP. Vertical transmission in chikungunya infection. Ceylon Med J. 2009;54:47–50.

33. Chua CL, Sam IC, Chiam CW, Chan YF. The neutralizing role of IgM during early chikungunya virus infection. PLoS One. 2017;12:e0171989.

34. Patil HP, Gosavi M, Kulkarni R, Mishra AC, Arankalle VA. Immunoglobulin G subclass response after chikungunya virus infection. Viral Immunol. 2022;35:437–42.

35. Griffin DE. 2013. Chapter 23 Alphaviruses. Knipe DM, Howley PM, eds. Fields virology, 6th ed, Philadelphia: Lippincott William & Wilkins. pp 651–686.

36. Jain J, Nayak K, Tanwar N, Gaind R, Gupta B, Shastri JS, et al. Clinical, serological, and virological analysis of 572 chikungunya patients from 2010 to 2013 in India. Clin Infect Dis. 2017;65:133–40.

37. Staples JE, Breiman RF, Powers AM. Chikungunya fever: an epidemiological review of a re-emerging infectious disease. Clin Infect Dis. 2009;49:942–8.

38. Bustos Carrillo F, Collado D, Sanchez N, Ojeda S, Lopez Mercado B, Burger-Calderon R, et al. Epidemiological evidence for lineage-specific differences in the risk of inapparent chikungunya virus infection. J Virol. 2019;93:e01622–18.

39. Gordon A, Gresh L, Ojeda S, Chowell G, Gonzalez K, Sanchez N, et al. Differences in transmission and disease severity between 2 successive waves of chikungunya. Clin Infect Dis. 2018;67:1760–7.

40. Kishishita N, Takeda N, Anuegoonpipat A, Anantapreecha S. Development of a pseudotyped-lentiviral-vector-based neutralization assay for chikungunya virus infection. J Clin Microbiol. 2013;51:1389–95.

41. Henss L, Yue C, Kandler J, Faddy HM, Simmons G, Panning M, et al. Establishment of an alphavirus-specific neutralization assay to distinguish infections with different members of the Semliki Forest complex. Viruses. 2019;11:82.

42. Su C, Ding K, Xu J, Wu J, Liu J, Shen J, et al. Preparation and application of chi pseudovirus containing double reporter genes. Sci Rep. 2022;12:9844.

43. Deng CL, Liu SQ, Zhou DG, Xu LL, Li XD, Zhang PT, et al. Development of neutralization assay using an eGFP chikungunya virus. Viruses. 2016;8:181.

44. Schmidt C, Perkovic M, Schnierle BS. Development of a sensitive detection method for alphaviruses and its use as a virus neutralization assay. Viruses. 2021;13:1191.

45. Pezzi L, LaBeaud AD, Reusken CB, Drexler JF, Vasilakis N, Diallo M, et al. GloPID-R chikungunya, o’nyong-nyong and Mayaro virus Working Group. GloPID-R report on chikungunya, o’nyong-nyong and Mayaro virus, part 2: Epidemiological distribution of o’nyong-nyong virus. Antiviral Res. 2019;172:104611.

46. Chanas AC, Hubalek Z, Johnson BK, Simpson DI. A comparative study of O’nyong nyong virus with chikungunya virus and plaque variants. Arch Virol. 1979;59:231–8.

47. Henss L, Yue C, Von Rhein C, Tschismarov R, Lewis-Ximenez LL, Dölle A, et al. Analysis of humoral immune responses in chikungunya virus (CHIKV)-infected patients and individuals vaccinated with a candidate CHIKV vaccine. J Infect Dis. 2020;221:1713–23.

48. Clements TL, Rossi CA, Irish AK, Kibuuka H, Eller LA, Robb ML, et al. Chikungunya and O’nyong-nyong viruses in Uganda: implications for diagnostics. Open Forum Infect Dis. 2019;6:ofz001.

49. LaBeaud AD, Banda T, Brichard J, Muchiri EM, Mungai PL, Mutuku FM, et al. High rates of o’nyong nyong and chikungunya virus transmission in coastal Kenya. PLoS Negl Trop Dis. 2015;9:e0003436.

50. Shah MM, Ndenga BA, Mutuku FM, Okuta V, Ronga CO, Chebii PK, et al. No Evidence of O’nyong-nyong viremia among children with febrile illness in Kenya (2015-2018). Am J Trop Med Hyg. 2021;104:1435–7.

