## Supplemental Table and Figure 1 for "Chikungunya virus antepartum transmission and abnormal infant outcomes in Nigeria"

**Supplemental Table 1.** Results of ChemBio CHIKV IgM/IgG rapid test for convalescent-phase samples from RT-PCR-confirmed ZIKV, DENV and CHIKV cases

| ChemBio DPP® IgM/IgG rapid test | No. of positive/total samples (%) in different serum/plasma panels* | | | | | |
|---|---|---|---|---|---|---|
| | negative | pDENV | sDENV | pZIKV | ZIKV wprDENV | CHIKV |
| CHIKV IgM† | 1/27 (3.7%) | 1/11 (9.1%) | 1/24 (4.2%) | 4/21 (19.0%) | 6/21 (28.6%) | **20/22 (90.9%)** |
| CHIKV IgG† | 0/27 (0%) | 0/11 (0%) | 0/24 (0%) | 4/21 (19.0%) | 6/21 (28.6%) | **22/22 (100%)** |

*pDENV, primary DENV infection; sDENV, secondary DENV infection; pZIKV, primary ZIKV infection; ZIKVwprDENV, ZIKV infection with previous DENV infection; CHIKV, CHIKV infection. Convalescent-phase serum/plasma samples were from RT-PCR-confirmed ZIKV, DENV and CHIKV cases as described previously and CHIKV-negative samples were from a dengue seroprevalence study in Taiwan (*20*-*22*); both CHIKV cases and CHIKV-negative samples were confirmed by CHIKV pseudovirus NT.

†The overall sensitivity/specificity of CHIKV IgM and IgG was 90.9/87.5% and 100/90.4%, respectively. The specificity of CHIKV IgM and IgG increased to 95.2% and 100%, respectively, when the pZIKV and ZIKVwprDENV panels with unknow history of CHIKV infection were not included in the calculation.

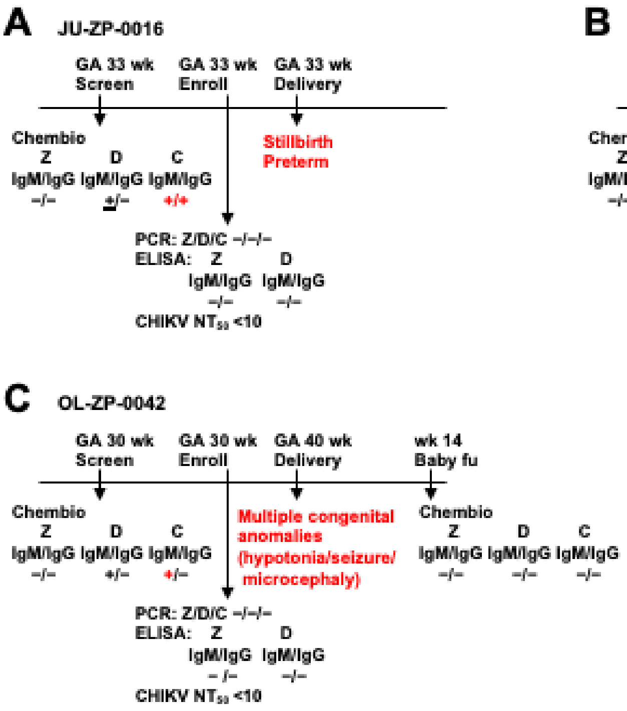


**Supplemental Figure 1.** Time course of 3 pregnant women with acute CHIKV infection not confirmed by NT and their infants with abnormal outcomes. Stillbirth and preterm (A), stillbirth (B), and multiple congenital anomalies (C).
